# Antigen testing for COVID-19 using image-based assessment of oral specimens

**DOI:** 10.1101/2022.05.27.22274752

**Authors:** Satoshi Shimozono, Mayu Sugiyama, Hiroshi Kurokawa, Hiroshi Hama, Masae Sato, Satoru Morikawa, Kumiko Kuwana, Kei Haga, Reiko Takai-Todaka, Shunsuke Inaura, Yuta Matsumura, Hidekazu Masaki, Naoto Nemoto, Ryoko Ando, Takako Kogure, Asako Tosaki, Hidehiro Fukuyama, Hideyuki Saya, Taneaki Nakagawa, Takuya Morimoto, Hiroshi Nishihara, Kazuhiko Katayama, Atsushi Miyawaki

**Affiliations:** Laboratory for Cell Function Dynamics, RIKEN Center for Brain Science, 2-1 Hirosawa, Wako-city, Saitama 351-0198, Japan; Keio Cancer Center, Keio University School of Medicine, 35 Shinanomachi, Shinjuku-ku, Tokyo 160-8582, Japan; Department of Dentistry and Oral Surgery, Keio University, School of Medicine, 35 Shinanomachi, Shinjuku-ku, Tokyo 160-8582, Japan; Department of Infection Control and Immunology, Ōmura Satoshi Memorial Institute, Kitasato University. 5-9-1 Shirokane, Minato-ku, Tokyo 108-8641, Japan; Safety Science Laboratories, Kao Corporation, 2-1-3 Bunka, Sumida-ku, Tokyo 131-8501, Japan; Epsilon Molecular Engineering Inc., Research bldg. of Open Innovation Center in Saitama University, 255 Shimo-Okubo, Sakura-ku, Saitama city, Saitama 338-8570, Japan; Biotechnological Optics Research Team, RIKEN Center for Advanced Photonics, 2-1 Hirosawa, Wako-city, Saitama 351-0198, Japan; Laboratory for Lymphocyte Differentiation, RIKEN Center for Integrative Medical Sciences, 1-7-22 Suehiro-cho, Tsurumi-ku, Yokohama City, Kanagawa 230-0045, Japan; Division of Gene Regulation, Institute for Advanced Medical Research, Keio University School of Medicine, 35 Shinanomachi, Shinjuku-ku, Tokyo 160-8582, Japan

## Abstract

While numerous diagnostic tests for COVID-19 have been developed for clinical and public health use, most of them provide binary or one-dimensional information on SARS-CoV-2 infection in pursuit of speed and ease of use. As their readouts are largely dependent on the specimen collection procedure, reliable diagnosis is still difficult. Here we report the development of a prototypical method for the immunocytochemical diagnosis of SARS-CoV-2 infection using oral specimens and fluorescent nanobodies against the viral spike and nucleocapsid proteins. Our cytological approach for the detection of SARS-CoV-2 infection was validated by our finding that at least half of SARS-CoV-2 RNAs in oral specimens were localized in the cellular fraction. Mapping antigens on sampled cells provided qualitative image data to which appropriate statistical texture analysis could be applied for the quantitative assessment of SARS-CoV-2 infectious status. A comprehensive comparative analysis revealed that oral cavity swabbing by medical workers provides specimens for COVID-19 diagnosis that yield comparable diagnostic accuracy as self-collected saliva specimens. Our diagnostic strategy may enable medical workers to acquire a wealth of information on virus–human cell interactions for multifaceted insight into COVID-19.

## Results

Since 2020, we have been developing techniques for visualizing SARS-CoV-2 by fusing fluorescent proteins (FPs) to recombinant antibodies that target the structural proteins of the virus, such as the S and N proteins. S is a surface spike (S) glycoprotein that is responsible for viral entry into the host cell. Over the past year, a number of potent neutralizing antibodies against the S protein have been developed. On the other hand, N is a nucleocapsid protein and is the most abundant structural protein in the virus. Many immunochromatographic tests for COVID-19 have been designed with the use of N as the target antigen. We employed nanobodies (Nbs), which are the variable domains of heavy chain–only antibodies (de Beer and Giepmans, 2020; Wrapp et al., 2020). Screening for S-binding ability in a cDNA library of 10 trillion synthetic Nb sequences yielded a high-affinity binder, K-874A Nb, that effectively alleviated symptoms in SARS-CoV-2–infected Syrian hamsters after nasal delivery (Haga et al., 2021). Since two recent studies successfully engineered multivalent Nb constructs with extremely high SARS-CoV-2 neutralization potency *in vitro* (Schoof et al., 2020; Xiang et al., 2020), we expected that multivalent Nb binding by fusion to a multimeric FP would enhance target binding by increasing avidity. Many of the wild-type FPs characterized so far form obligate multimers (Miyawaki, 2011). We took interest in a green-emitting FP that we previously cloned from the stony coral *Favia favus* (Tsutsui et al., 2005). The FP, called KikG, forms an obligate tetrameric complex.

We designed functional fluorescent Nbs by combining K-874A and KikG. One of the constructs, K-874A=KikG, has a coupler linker (designated by =) (Shimozono and Miyawaki, 2008) between the two domains. This design resulted in good yield, soluble expression in bacterial culture, and efficient purification of the recombinant chimeric protein. The full maturation of the KikG chromophore and the molecular integrity of the fusion construct were verified by absorption measurement and SDS-PAGE, respectively (Fig. S1).

VeroE6 cells constitutively expressing TMPRSS2 were infected with a strain of SARS-CoV-2 (KUH003) at a multiplicity of infection of 0.1, fixed at 24 h post-infection, and processed for immunoreaction with K-874A=KikG. Green fluorescent puncta were clearly observed in the infected cells that reacted with K-874A=KikG, but not in uninfected cells or non-reacted infected cells, indicating that the signal was specific to SARS-CoV-2 infection and the S protein (Fig. 1a, left). Our comparative experiments using different FPs revealed that K-874A=KikG produced much stronger fluorescence signals than any other constructs (Fig. 1a, right, Fig. S1); when KikG was replaced with EGFP or Achilles, the latter of which is a fast-maturing, yellow-emitting FP, only a faint signal was observed under the same optical conditions. It should be noted that both EGFP and Achilles were assumed to be virtually monomeric. Substituting the red-emitting FP AzaleaB5, which we previously cloned (Ando et al., 2020) and which forms an obligate dimeric complex, led to the generation of substantial signals with a conventional red fluorescent protein filter set (Fig. 1b, Fig. S1). Taken together, these findings confirmed that the multimeric nature of FP is critical for the efficient detection of the trimeric S protein complex.

**Fig. 1.**
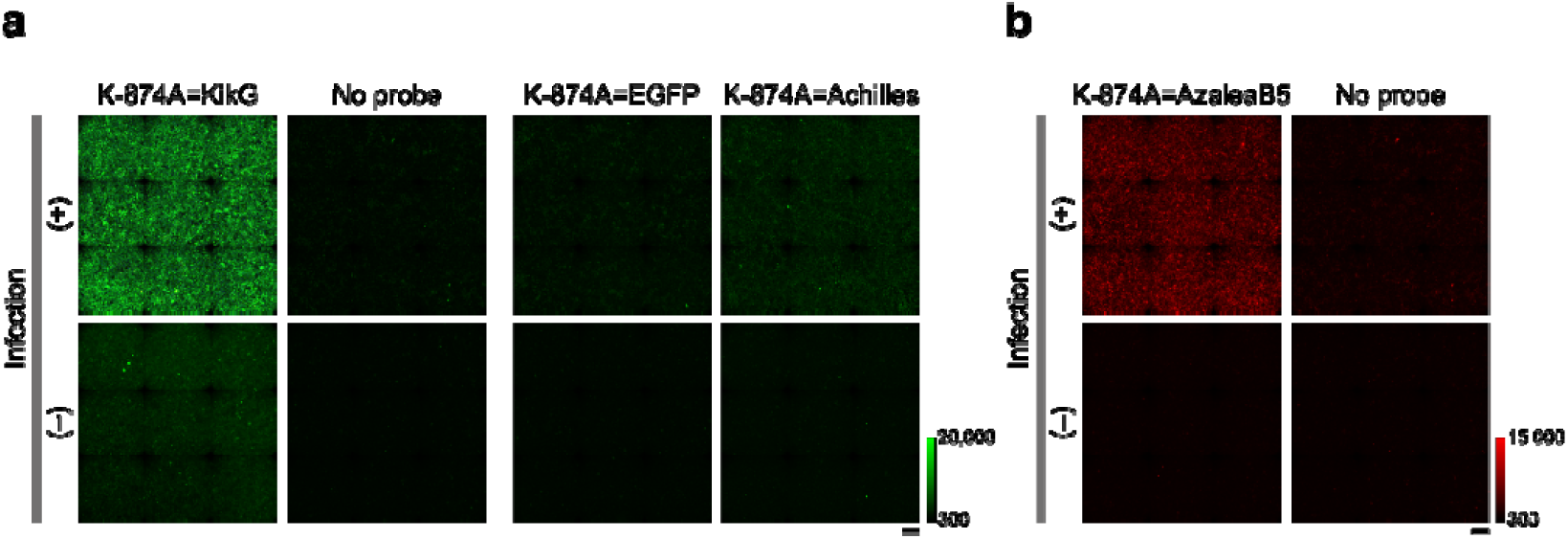
Wide-field microscopic images of VeroE6/TMPRSS2 cells infected with a strain of SARS-CoV-2, KUH003. Infected (top) and uninfected (bottom) cells were treated or not treated with anti–S protein Nb (K-874A) fused to FPs. “=” denotes “coupler linker,” a triple repeat of the amino acid linker Gly–Gly–Gly–Gly–Ser used for fusion. Full cell confluency in each imaged area was confirmed by DAPI staining. The green or red scale indicates the lowest and highest image intensities. Scale bars, 0.2 mm. (a) A U-FBNA filter cube (excitation: 470–495, dichroic mirror: 505, emission: 510–550, Olympus) was used to observe KikG, EGFP, and Achilles fluorescence. (b) A U-FGNA filter cube (excitation: 540–550, dichroic mirror: 570, emission: 575–625, Olympus) was used to observe AzaleaB5 fluorescence.

SARS-CoV-2 is constantly mutating, and variants have emerged and circulated worldwide over the past year. These variants are mostly defined by multiple mutations in the S protein. We prepared VeroE6/TMPRSS2 cells infected not only with KUH003 but also with the Alpha, Beta, or Gamma variants, and examined the immunoreactivity of K-874A=KikG toward these infected cells (Fig. 2, top). We found that the Gamma variant was a less effective target of K-874A=KikG than the other variants. Accordingly, we screened the aforementioned library for Nbs with improved S protein binding. Our *in vitro* experiments indicated that one of the Nbs, namely E9, was generous to mutations in the S protein. We replaced K-874A with E9 to generate E9=KikG, and verified that the biochemical and spectroscopic properties of this new reagent were as good as those of K-874A=KikG (Fig. S1). We found that E9=KikG equally detected all the SARS-CoV-2 variants examined in this study, and generated stronger signals than K-874A=KikG (Fig. 2, bottom). Thereafter, we used E9=KikG to visualize the S protein.

**Fig. 2.**
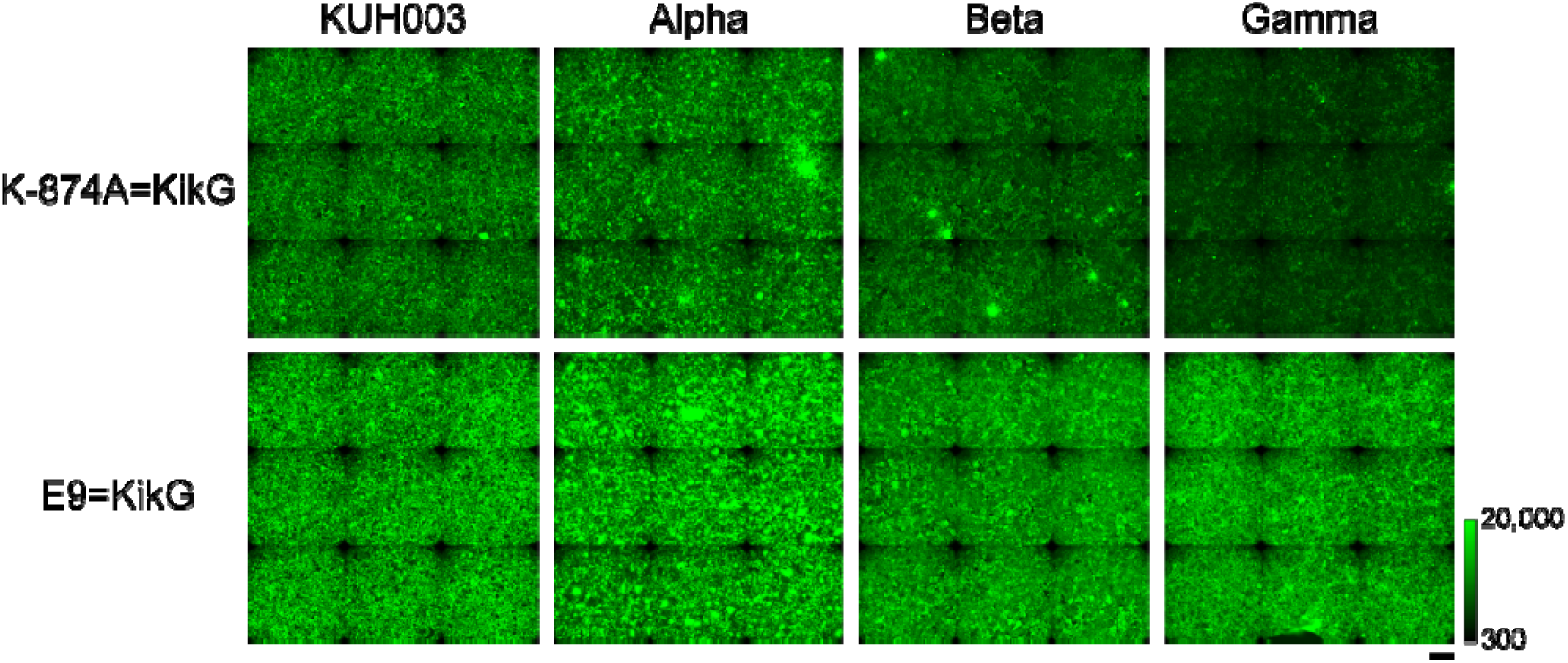
Comparison of SARS-CoV-2 variant detection spectra between K-874A=KikG (top) and E9=KikG (bottom). VeroE6/TMPRSS2 cells were infected with KUH003, Alpha, Beta, or Gamma variants. Full cell confluency in each imaged area was confirmed by DAPI staining. Fluorescence images were acquired by wide-field microscopy. The green scale indicates the lowest and highest image intensities. Scale bar, 0.2 mm.

We applied E9=KikG to a diagnostic test for COVID-19. An exhaustive study by Huang et al. (2021) demonstrated that the oral cavity is an important site for SARS-CoV-2 infection. In the present study, therefore, we used oral specimens for the preparation of smears (Fig. 3a). A participant was asked to spit saliva into a collection bottle; hereinafter, this self-collected saliva is abbreviated as “SS.” An SS specimen (#DP29) taken from a symptomatic COVID-19 inpatient (DP29) had a relatively high viral load with a cycle threshold (Ct) value of 29.963, and was used as a positive control. By contrast, an SS specimen (#IC2) taken from a healthy person (IC2) proved to be useful as a negative control (Ct > 40). These collected specimens were first placed in preservative medium used in the liquid-based cytology (LBC) method, which has been widely employed for routine screening of cervical lesions. Then, smears of the two specimens were made on silane-coated slides for immunostaining with E9=KikG and for nuclear staining with DAPI. We used conventional wide-field microscopy for observation. Green fluorescence from KikG was abundantly detected in the cell populations from #DP29 but not in the cell-free regions or the cells from #IC2 (Fig. 3b, top). These results suggested that this method using E9=KikG resulted in a high signal-to-noise ratio (SNR).

**Fig. 3.**
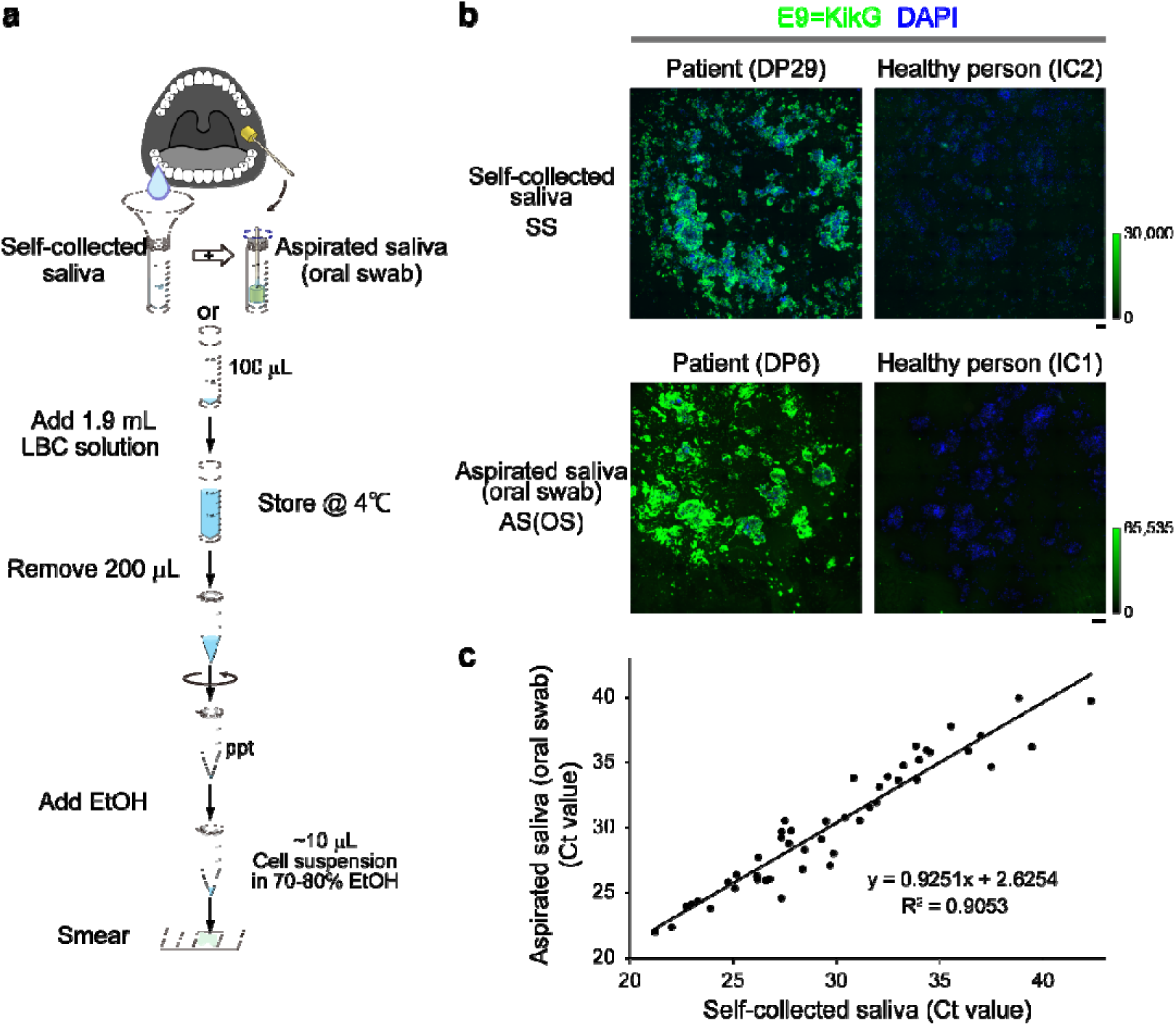
Diagnostic tests for COVID-19 based on immunoreaction of E9=KikG using oral specimens. Fluorescence images were acquired by wide-field microscopy. (a) Schematic of the LBC method for preparing smears from SS and AS(OS) specimens. LBC: liquid-based cytology. (b) *top*, Immunosignals (green) of E9=KikG in SS specimens from a symptomatic COVID-19 patient (DP29) and a healthy person (IC2). *bottom*, Immunosignals (green) of E9=KikG in AS(OS) specimens from a symptomatic COVID-19 patient (DP6) and a healthy person (IC1). Nuclei were counterstained with DAPI (blue). Scale bars, 0.2 mm. (c) Pairwise association of Ct values between AS(OS) and SS specimens. Simple regression analysis revealed a strong correlation, with a coefficient of determination (R^2^) of 0.9053. The equation for the regression line in which *y* and *x* were defined as the Ct values of AS(OS) and SS specimens, respectively, was *y* = 0.9251 *x* + 2.6254, suggesting near equivalence for the detection of SARS-CoV-2 RNAs between the two specimens.

In another set of experiments, saliva was aspirated from the oral cavity into a collection bottle. Furthermore, a swab was rotated several times against the tongue, buccal mucosa, and gingiva and was placed into the collection bottle. Hereinafter, the combination of aspirated saliva and oral swab is abbreviated “AS(OS).” AS(OS) specimens were prepared from a symptomatic COVID-19 inpatient (DP6) and a healthy person (IC1) (Fig. 3a). We found that E9=KikG identified SARS-CoV-2 infection in the AS(OS) specimen (#DP6, Ct value: 29.665), again with a high SNR (Fig. 3b, bottom).

In general, researchers have immunologically identified SARS-CoV-2 infection by employing an anti–SARS-CoV-2 S protein monoclonal antibody (mAb) (1A9) (GeneTex, GTX632604) and a fluorescently labeled secondary antibody. However, when we used this de facto standard method, we noticed a substantial level of fluorescence in regions devoid of cells (Fig. S2). This background fluorescence appeared to come from 1A9 binding, because it was not apparent with the secondary antibody itself (data not shown).

To directly compare the diagnostic sensitivity achieved using SS and AS(OS) specimens, in separate experiments we quantitated the SARS-CoV-2 RNA copies (Ct values) of the two specimens that were collected simultaneously from each of 25 inpatients with COVID-19. The two specimens showed comparable Ct values in most cases (Fig. 3c).

Although our study adopted a cytological approach that focused on cellular fractions, it was important to know the extent to which we ignored the free viruses present in acellular fractions. According to the study by Huang et al. (2021), both acellular and cellular salivary fractions from COVID-19 patients exhibited a substantial level of infectivity. As illustrated in Fig. 4a, we attempted to roughly estimate the viral quantity ratio between the two fractions derived from 12 specimens (four SS and eight AS(OS)) collected from COVID-19 patients. We centrifuged each specimen to prepare a supernatant and a pellet, which corresponded to the acellular and cellular fractions, respectively. The two fractions were treated with proteinase and diluted with phosphate-buffered saline (PBS) to the same volume for the parallel quantification of viral RNA genomes by RT-qPCR. The Ct value of the cellular fraction was equal to or slightly lower than that of the acellular fraction (Fig. 4b). Based on this result, we postulated that over half of the viruses in the original specimen were concentrated in the cell preparations. We thus reasoned that methods of cytological diagnosis, including LBC, are able to sensitively detect SARS-CoV-2 infection because cells can be easily collected by sedimentation or centrifugation, and because infected cells containing abundant SARS-CoV-2 viruses can be identified by immunofluorescence techniques with high SNRs.

**Fig. 4.**
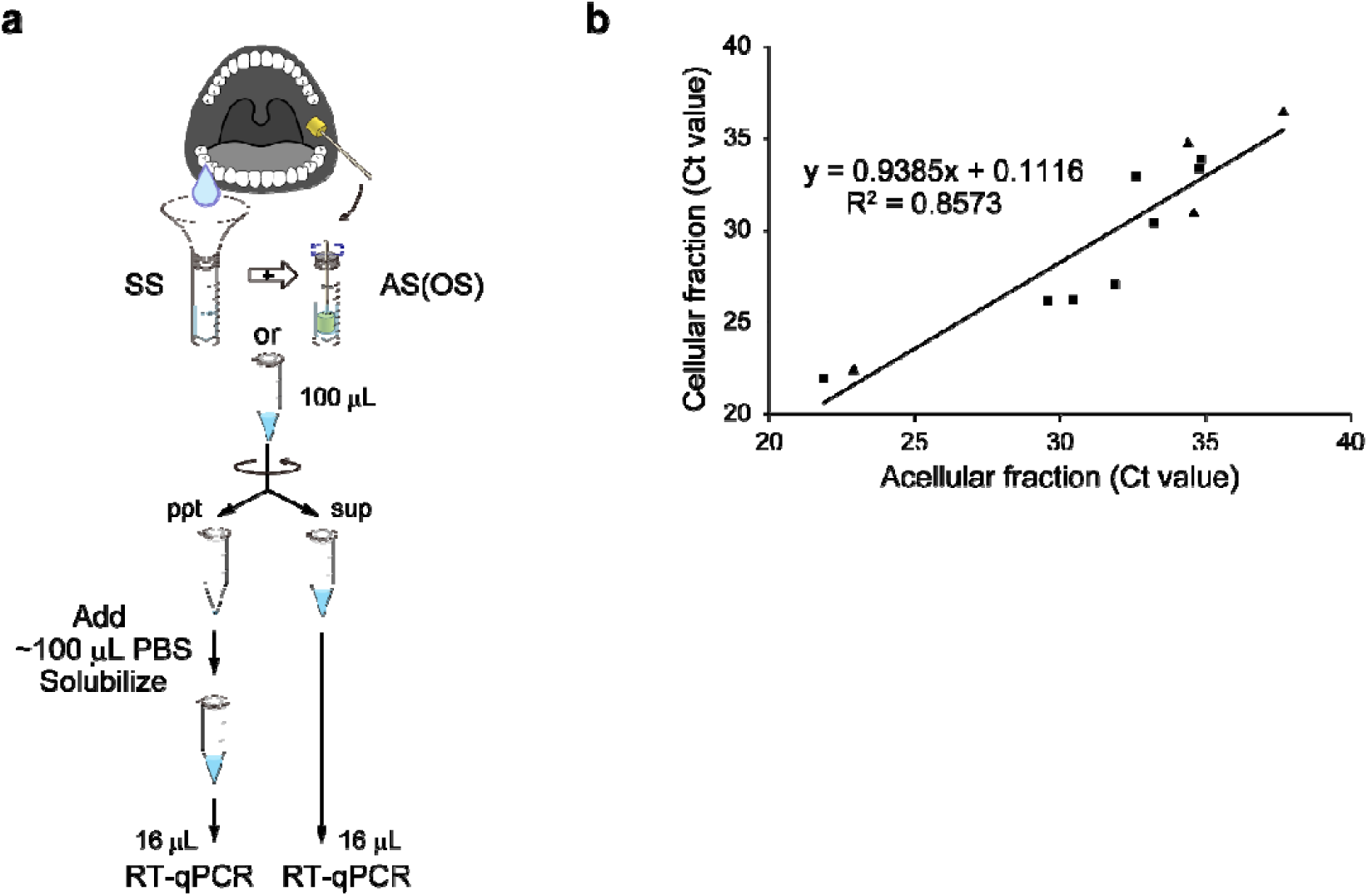
Estimation of SARS-CoV-2 RNA quantity in cellular fractions from SS and AS(OS) specimens. (a) Schematic of the work-in-process for parallel RT-qPCR. PBS: phosphate-buffered saline. (b) Pairwise association of Ct values between cellular and acellular fractions. Simple regression analysis revealed a strong correlation between them, with a coefficient of determination (R^2^) of 0.8573. The equation for the regression line in which *y* and *x* were defined as the Ct values of cellular and acellular fractions, respectively, was *y* = 0.9385 *x* + 0.1116, suggesting nearly equal distribution of SARS-CoV-2 RNAs into the pellet and the supernatant. AS(OS) specimens: solid triangles. SS specimens: solid squares.

However, we noticed a certain degree of deterioration of cellular morphology of the collected cells after several week storage in the LBC medium and shipment. We therefore designed an alternative procedure that employed paraformaldehyde (PFA) for the reliable fixation of cells collected from the oral cavity (Fig. 5). After fixation with 4% PFA and washing with PBS, cellular fractions obtained by centrifugation were suspended in 70–80% ethanol for smear preparation. A suspension derived from a tiny amount (∼10 μL) of the original specimen was spread on a silane-coated slide. The specimen was subjected to cytological analysis that involved SARS-CoV-2 detection by E9=KikG and counterstaining of nuclei with DAPI. High-resolution imaging was performed using a laser-scanning confocal microscope (Olympus, FV3000) equipped with a high numerical aperture (NA) objective lens (Olympus, UPLFLN-40XO 40×/1.30 NA). At each *xy* position, multiple confocal images were acquired at different *z* positions (*z* step: 1.5 μm) to generate a *z*-stack image. A large number (∼1,000) of *z*-stack images were generated at neighboring *xy* positions and combined using the microscope system’s “tiling” software.

**Fig. 5.**
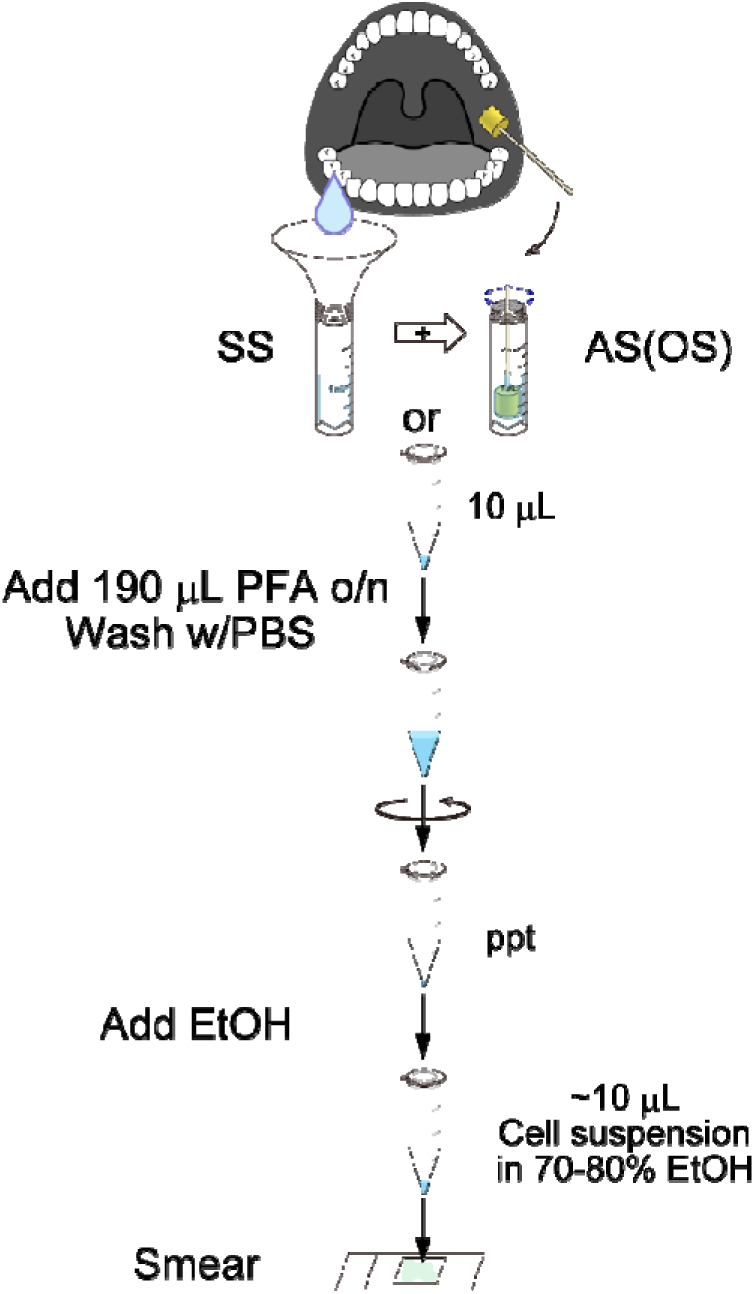
Schematic of smear preparation for comprehensive, high-resolution imaging of SARS-CoV-2 in cellular fractions from AS(OS) and SS specimens.

In this way, we generated exhaustive dual-color image data for the cells present in 7.5 μL of an AS(OS) specimen (#DP118-1_AS(OS)) (Fig. S3) taken from a COVID-19 inpatient (DP118) in the early stage of infection. This AS(OS) specimen had a high viral load (Ct value: 22.002). This comprehensive imaging approach allowed us to categorize the pattern of E9=KikG signals into three groups. First, quite a few uninfected cells showed no signals (Fig. 6a, arrows). Second, a fraction of cells exhibited strong, homogeneous E9=KikG fluorescence labeling (Fig. 6a, solid arrowheads). These infected cells probably contained a large number of viruses. Third, many cells were characterized by a spotty or circumferential pattern of green fluorescence (Fig. 6a, open arrowheads). It is possible that these cells had already discharged most viral particles and would soon die. After death, the membrane structures containing the S protein would become entangled to form insoluble materials. In fact, we occasionally noticed cell debris that was strongly labeled with E9=KikG. Importantly, all these E9=KikG signals were specific to SARS-CoV-2 infection; they were never seen in the specimens from healthy persons. This justifies our cytological approach as a quantitative method.

We prepared an SS specimen, #DP118-1_SS, on the same day that #DP118-1_AS(OS) was obtained, and it too had a high viral load (Ct value: 21.238). We performed the same cytological analysis on #DP118-1_SS. The green signal of E9=KikG was quite stable; it did not fade or become blurred at all 3 days after slide preparation (encapsulation) (Fig. S4). This signal stability allowed us to compare new smears with old ones across days. The aforementioned inpatient (DP118) provided another pair of specimens, #DP118-2_AS(OS) and #DP118-2_SS, 6 days after the first pair (#DP118-1_AS(OS) and #DP118-1_SS). #DP118-2_AS(OS) and #DP118-2_SS showed relatively high Ct values (36.275 and 33.833, respectively), suggesting that the patient was in remission. To investigate how the three types of cell populations (Fig. 6a) changed over time, we performed quantitative analysis on the four specimens. The analysis consisted of segmentation, feature extraction, and k-means clustering (Mohapatra et al., 2011). After manual segmentation, two features of the E9=KikG fluorescence were automatically extracted from individual cells. One was the average fluorescence intensity, which was calculated straightforwardly. The other was texture, which was defined as a function of the spatial variation in pixel intensities and was used to effectively highlight the spotty and circumferential patterns. Our texture analysis was performed in three steps. First, we constructed fluorescence intensity profiles along both vertical and horizontal lines throughout each segmented cell. Second, we carried out fast Fourier transform analysis on each line plot with 64 (2^6^) divisions. Third, we quantified the strength of the high-frequency components for each cell and created 2D scatter plots by plotting the frequency against the average intensity for all segmented cells (n = 300, 304, 121, and 300 for #DP118-1_AS(OS), #DP118-2_AS(OS), #DP118-1_SS, and #DP118-2_SS, respectively) (Figs. 6b and 6c, top). Finally, we used k-means to classify the labeling of the cells as strong, sparse, or none. Although the percentage of infected cells was greatly reduced in the late stage of infection, the ratio of strong to sparse labeling remained nearly the same (Figs. 6b and 6c, bottom).

**Fig. 6.**
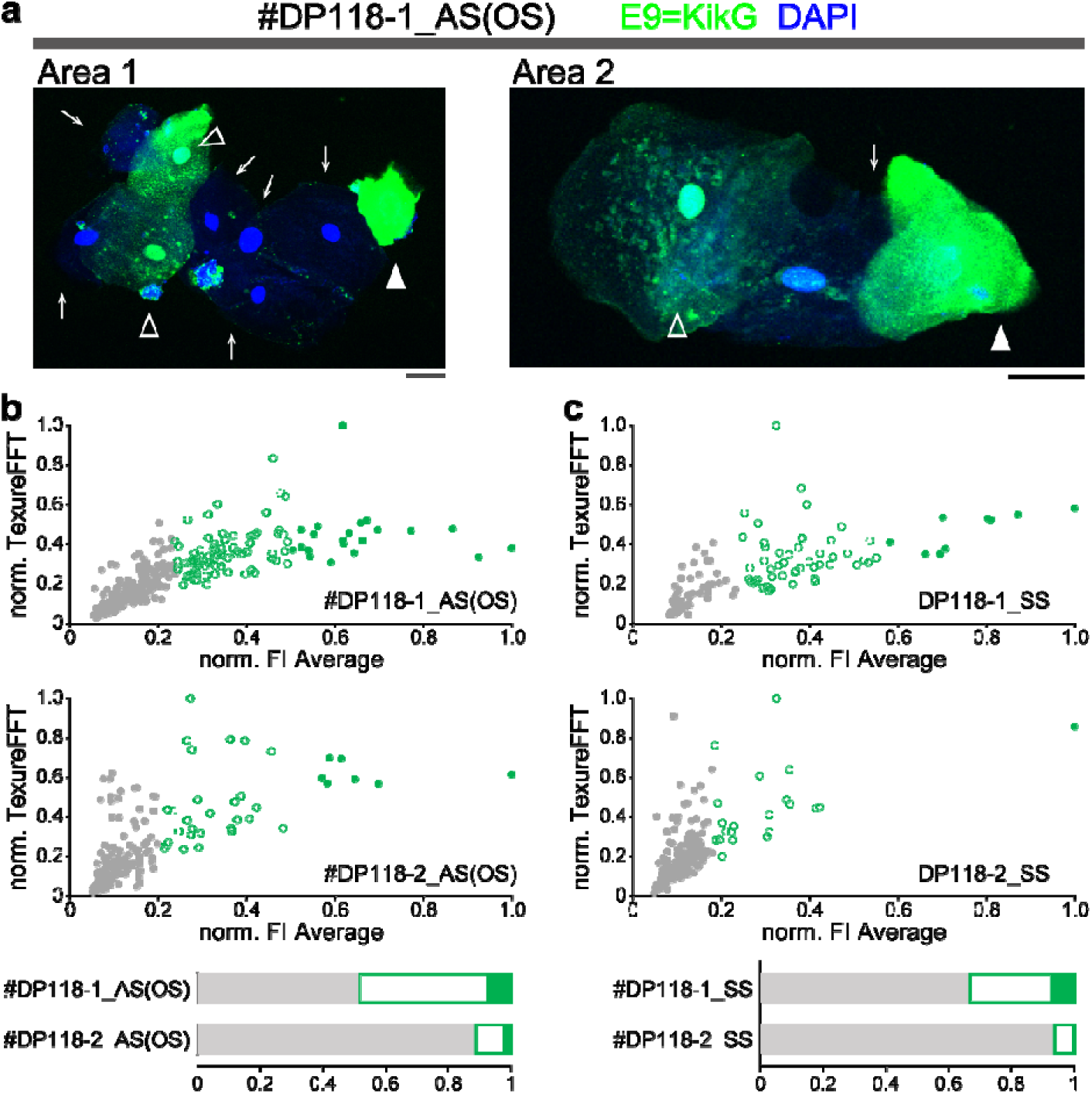
Comprehensive, high-resolution confocal imaging of SARS-CoV-2 in cellular fractions using E9=KikG. (a) Three types of E9=KikG immunosignals (green) identified in an AS(OS) specimen (#DP118-1_AS(OS)) from a COVID-19 inpatient (DP118) in the early stage of infection. Nuclei were counterstained with DAPI (blue). Two representative images (Areas 1 and 2) are illustrated. Cells with strong, sparse, and no labeling are indicated by solid arrowheads, open arrowheads, and arrows, respectively. Scale bars, 0.2 mm. In the sparsely labeled cells, spotty fluorescence was sometimes observed on the nucleus. (b) Analysis of two AS(OS) specimens from patient DP118: #DP118-1_AS(OS) and #DP118-2_AS(OS). (c) Analysis of two SS specimens from patient DP118: #DP118-1_SS and #DP118-2_SS. (b, c) top, Scatter plots of averaged fluorescence intensity (FI) vs. texture (FFT). Data points of cells with strong, sparse, and no labeling with E9=KikG are indicated by solid green, open green, and solid gray circles, respectively. Bottom, bar charts represent the relative population size of cells with strong (solid green), sparse (open green), and no (solid gray) labeling.

It should be noted that the smears in this study sometimes contained thick epithelial fragments. Whereas such materials are morphologically undiagnosable and thus usually avoided in the conventional LBC method, we adopted a simple yet comprehensive approach that effectively detected the E9=KikG fluorescence, even in thick fragments. Moreover, whereas the LBC method eliminates interfering materials that obscure diagnosis, such as erythrocytes and leukocytes, our approach was able to target non-epithelial cells as well. In a separate experiment that used E9=KikG and DAPI, we visualized squamous epithelial cells in oral specimens using an antibody against pan-cytokeratin (pCK). As also reported by Huang et al. (2021), we noticed that a significant number of pCK-negative cells, probably inflammatory cells, exhibited E9=KikG fluorescence. Such pCK-negative infected cells were observed in both AS(OS) and SS specimens (Fig. 7).

**Fig. 7.**
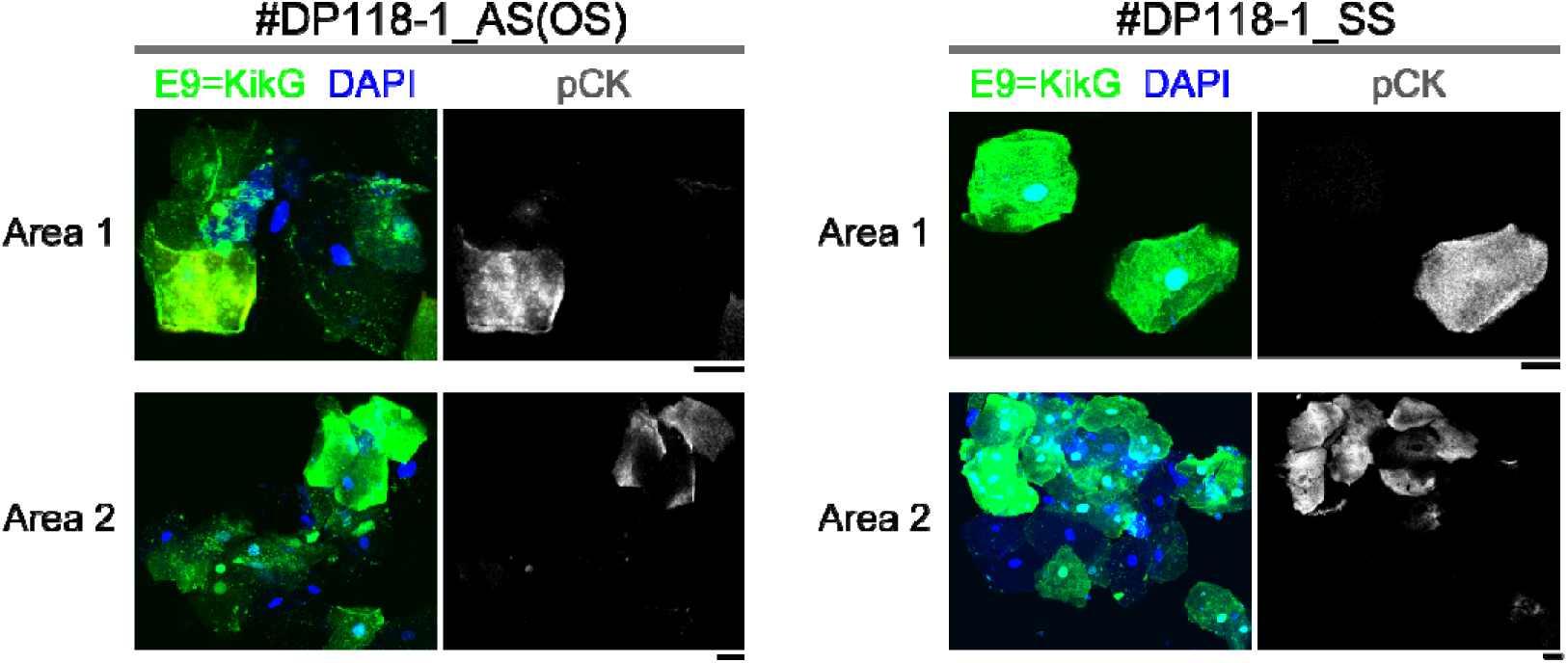
Epithelial and non-epithelial SARS-CoV-2–infected cells observed in an AS(OS) specimen (#DP118-1_AS(OS), Ct = 22.002) and an SS specimen (#DP118-1_SS, Ct = 21.238). Two representative confocal images are shown for each specimen. pCK: pan-cytokeratin. Scale bars, 0.2 mm.

We were interested in the fact that most of the SARS-CoV-2–infected cells expressed the S protein on the plasma membrane. These cells might be allowed to fuse with other cells that expressed the ACE2 receptor protein. Braga et al. (2021) recently characterized SARS-CoV-2 S–induced syncytia in their post-mortem histologic analysis of COVID-19 patient lungs; most of the syncytia were identified as pneumocytes. Since the study by Huang et al. (2021) revealed that multiple oral epithelial cell subtypes expressed ACE2, we wondered if multinucleated syncytia would be detectable in oral specimens. We used the #DP118-1_SS saliva specimen. To clearly outline individual epithelial cells, we immunostained the plasma membrane using anti–pan-cadherin antibody and were able to identify syncytia that had a large cytoplasm containing multiple nuclei (Fig. 8).

**Fig. 8.**
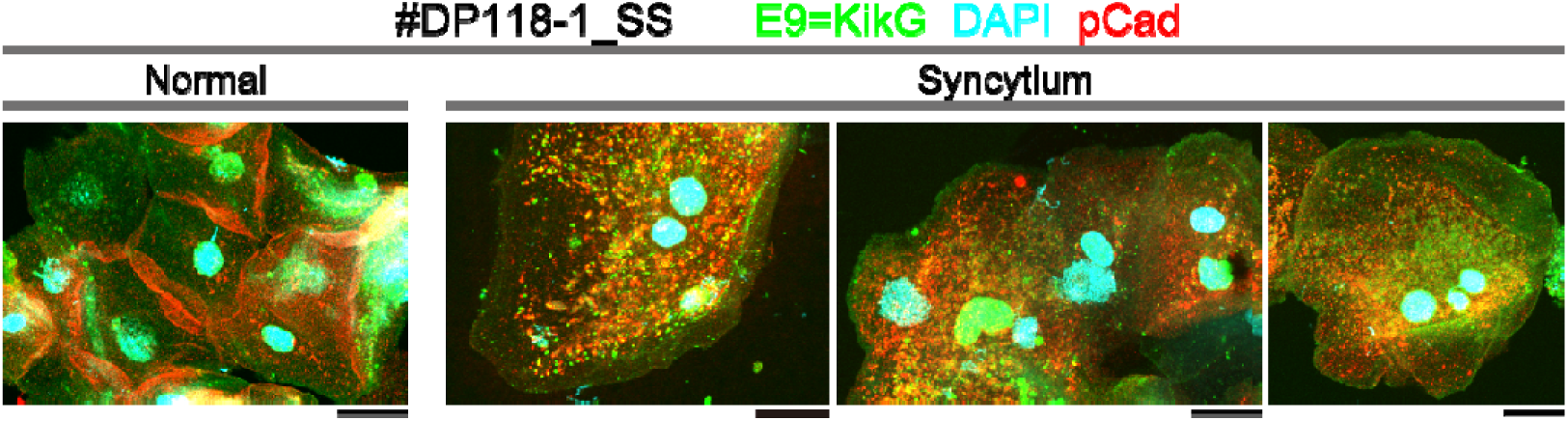
Syncytium formation in SARS-CoV-2–infected epithelial cells observed in an SS specimen (#DP118-1_SS, Ct = 21.238). Three representative confocal images of cells with syncytia are shown. pCad: pan-cadherin. Scale bars, 0.2 mm. Syncytia were also found in an AS(OS) specimen (data not shown).

Sherwood and Hayhurst (2021) generated fluorescent immunoreagents by fusing a new Nb against the SARS-CoV-2 N protein to mNeonGreen and mScarlet-I, and characterized their immunoreactivity toward SARS-CoV-2–infected Vero cells. As such, we screened for SARS-CoV-2 N–binding ability in the cDNA library to obtain high-affinity Nbs. One of them, N10 Nb, was successfully fused to AzaleaB5 via the coupler linker to generate N10=AzaleaB5 (Fig. S1). This red fluorescent Nb was found to detect SARS-CoV-2 infection (Fig. 9a). Although N is much more abundant in the virus than S, the fluorescence intensity of N10=AzaleaB5 was somehow weaker than that of E9=KikG. We exchanged FPs between the two constructs to generate N10=KikG and E9=AzaleaB5 (Fig. S1). This combination gave rise to a better signal balance between the green and red channels, and their merged signal (yellow) was expected to indicate the presence of viral particles and debris in the cellular fractions (Fig. 9b) more reliably than the E9=KikG signal alone.

**Fig. 9.**
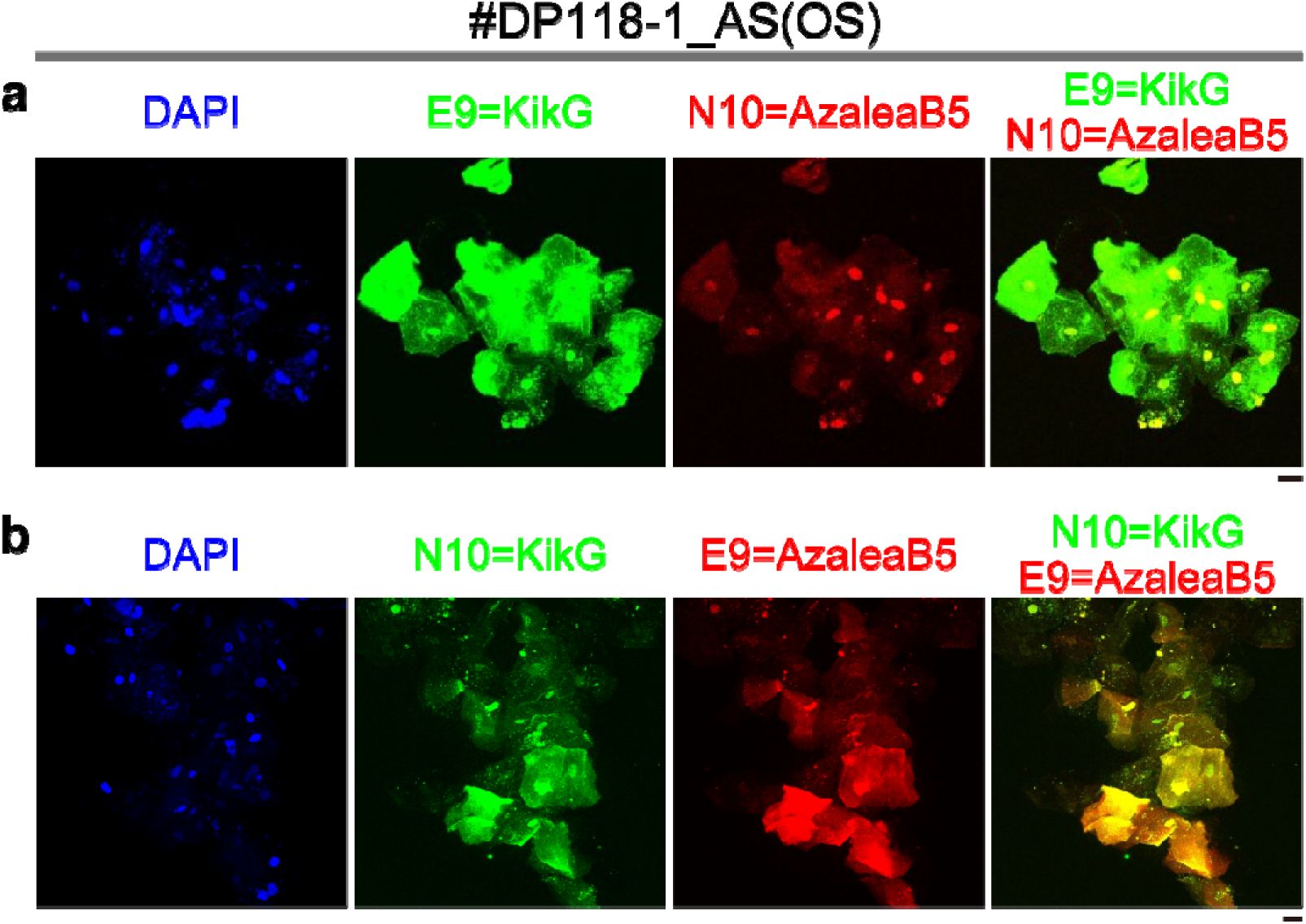
Immunostaining for both S and N proteins in SARS-CoV-2–infected cells in the #DP118-1_AS(OS) specimen. After fixation with 4% PFA, smears were treated with E9=KikG and N10=AzaleaB5 (a) or N10=KikG and E9=AzaleaB5 (b). Nuclei were counterstained with DAPI (blue). Representative confocal images are shown. Scale bars, 0.2 mm. The merged green and red images are shown in the rightmost column.

## Discussion

Currently, common antigen tests detect SARS-CoV-2 proteins through directional flow. Some of the most recent tests no longer require complex instruments or skilled operators. Due to their low sensitivity, however, they still require larger amounts of viral material than PCR-based tests. By contrast, cytological approaches to antigen testing may enable reliable diagnosis of COVID-19. Our comparative RT-qPCR experiments (Fig. 4) revealed that SARS-CoV-2 viruses were abundant in the cellular fractions of oral specimens from COVID-19 patients. Our cytological experiments revealed that infected cells were detectable even in specimens taken from high-Ct patients. Importantly, only a few microliters of oral cavity–derived fluid were sufficient for diagnosing COVID-19 through the direct visualization of SARS-CoV-2, indicating that the problem of low sensitivity inherent to common antigen tests is not an issue for our cytological approach coupled with image-based assessment. Cytological examination may require medical workers at the point of care to have some expertise and experience, but the tasks involved will soon be automated through the implementation of artificial intelligence technologies. Our method will be performed in central laboratories with specialized equipment including imaging flow cytometry, which acquires multichannel images of a large number of single cells within minutes (Doan et al., 2018).

A previous study performed liquid-based immunocytochemical detection of SARS-CoV-2–infected squamous cells in clinical nasopharyngeal swab (NPS) specimens using a primary antibody against the SARS-CoV-2 N protein and a reporter-coupled secondary antibody (Parada et al., 2021). By contrast, we employed Nbs, which can be genetically engineered to be fluorescent via FP fusions. Our method does not require a secondary antibody, which considerably decreases the time required for the immunocytochemical detection of SARS-CoV-2 infection. It should be noted that Nb-FP fusion proteins can be mass produced in bacteria. When E9=KikG was expressed in *Escherichia coli* and purified, for example, the soluble fraction contained approximately 80 mg of spectrally active proteins per liter of culture. Furthermore, we demonstrated that the ability of E9=KikG to detect SARS-CoV-2 infection was unaffected by lyophilization (Fig. S5). Thus, the transportation and storage of the assay reagent would require neither cooling nor freezing, a boon to clinical laboratories in remote areas. Taken together, these findings imply that the cytological approach employing our fluorescent Nbs could be used globally for rapid and cost-effective diagnostic testing of COVID-19, complementing PCR and immunochromatography. In the future, our approach may be effectively coupled with a mobile phone–based fluorescence microscope for the purpose of both personal and population health.

SARS-CoV-2 continuously evolves by means of genetic mutations, mainly in the spike protein. The emergence of new variants has attracted global attention in the past year, as Delta and then Omicron were classified as new variants of concern (VOCs). We preliminarily noticed the ability of E9=KikG to react with infected cells in oral specimens from Delta patients. In our future research it will be important to examine if E9=KikG can be used for the practical diagnosis of Delta or Omicron infection, and to further evolve the Nb toward these new VOCs.

NPSs and oro-nasopharyngeal swabs (ONPSs) are used as standard specimens for COVID-19 diagnosis. However, these procedures are quite invasive and may stimulate sneezing and coughing, posing significant risks to frontline medical workers. Sampling in the oral cavity may be an alternative to NPS and ONPS. In particular, saliva can be easily self-collected and does not require swabs. Some literature (Lee et al., 2021; Torretta et al., 2021) has shown that SS specimens may be an acceptable alternative source for the diagnosis of SARS-CoV-2 infection by RT-qPCR. On the other hand, the diagnostic potential of oral swab specimens has not yet been explored. Oral swabbing would be particularly effective for patients who have a high fever and cannot spit saliva due to dehydration. Although oral swabbing could also be performed by patients who are given appropriate instructions, oral swab collection by medical workers will still be essential for diagnosing gravely ill patients who are either unconscious or who present with dementia or motor disability. Remarkably, our comparison of paired SS and AS(OS) specimens revealed high similarity between the cell populations from the two types of specimens in terms of both immunoreactivity for SARS-CoV-2 and viral RNA copy numbers (Ct values) (Fig. 3c). Accordingly, we conclude that SS and AS(OS) specimens are comparable materials for COVID-19 diagnosis.

Transmission electron microscopy data have revealed that a single infected cell contains as many as 10^5^ viral particles (Sender et al., 2020). It is likely that infected cells in expelled saliva droplets contribute to the efficient airborne transmission of SARS-CoV-2. Our imaging data showing that saliva droplets from COVID-19 patients are effectively SARS-CoV-2 “cluster bombs” may be sufficiently remarkable to convince the general public of the necessity to wear masks. The microscopic identification of strongly immunolabeled infected cells should be able to provide warning signs. Indeed, our approach can be used to effectively screen for COVID-19 super-spreaders.

## Data Availability

All data produced in the present study are available upon reasonable request to the authors.

## Acknowledgments

The authors thank Dr. H. Matsumoto (the president of RIKEN), Dr. S. Koyasu and Dr. K. Yamamoto at RIKEN, and Dr. M. Amagai at Keio University Hospital for special support, Drs. K. Midorikawa and A. Nakano at RIKEN Center for Advanced Photonics for general support, K. Higuchi and Y. Ue at RIKEN CBS-Olympus Collaboration Center and K. Mochida at Keio Cancer Center for technical assistance, Drs. Y. Uwamino, K. Koyama and M. Murata at Keio University Hospital for valuable support. This work was supported in part by RIKEN President’s Discretionary Fund (to A. M.); the COVID-19 Kitasato project (to K. Katayama); grants from the Japan Ministry of Education, Culture, Sports, Science and Technology Grant-in-Aid for Scientific Research on Innovative Areas: ‘Resonance Bio’ (15H05948 to A. M.); Platform Project for Supporting Drug Discovery and Life Science Research (Basis for Supporting Innovative Drug Discovery and Life Science Research), Japan Agency for Medical Research and Development (AMED) (20he1422004j to H. N. and 21ae0121001 to K. Katayama).

## Author Contributions

S. S. and M. S. developed Nb-FP fusion technology. S. S., M. S., H. K., and H. H. performed fluorescence imaging experiments and analyzed image data. H. K. devised the algorithm for image analysis. S. M. and T. N. collected clinical samples. R. A. and T. K. performed low magnification imaging experiments. T. M., H. M., N. N., Y. M., K. H., R. T.-T., and K. Katayama developed Nbs. K. H., R. T.-T. and K. Katayama performed infection experiments. S. I. and T. M. analyzed the performance of Nb-FP fusions. A. T. conducted lyophilization. H. F. and H. S. established a research system for clinical studies. S. M., M. S., K. Kuwana, T. N., and H. N. prepared clinical specimens for microscopy observation. M. S., K. Kuwana, and H. N. performed molecular diagnostic tests for COVID-19. H. N. supervised clinical studies. K. Katayama supervised infection experiments and Nb development. A. M. conceived the whole study, wrote the manuscript, and supervised the project.

## Competing interests

S. S., M. S., H. H., K. H., R. T.-T., S. I., T. M., K. Katayama, and A. M. are inventors on patent application that covers the creation and use of the VHHs (E9 and N10) fused with fluorescent proteins.

2021-166259

## Methods

### Protein purification

Recombinant proteins with a polyhistidine tag at the N-terminus were expressed in *Escherichia coli* [JM109 (DE3)]. Transformed *E. coli* was incubated in a Luria-Bertani (LB) medium containing 0.1 mg/mL ampicillin at room temperature (RT) with gentle shaking for several days. Protein purification by Ni^2+^ affinity chromatography was performed as described previously (Ando et al., 2002).

### In vitro spectroscopy

Absorption spectra were acquired using a spectrophotometer (U-3310, Hitachi). Fluorescence excitation and emission spectra were acquired using a fluorescence spectrophotometer (F-4500, Hitachi). Protein concentrations were measured using a Bradford assay kit (Bio-Rad) with bovine serum albumin (BSA) as the standard.

### SDS-PAGE

Proteins were denatured by boiling in sample buffer containing β-mercaptoethanol and then electrophoresed with 10% (wt/vol) polyacrylamide gel containing SDS according to Laemmli’s method. A prestained 10-kD protein ladder (Thermo Fisher/Invitrogen) was used as a size marker. Proteins were detected by staining with Coomassie Brilliant Blue.

### cDNA display for obtaining Nbs E9 and N10

Preparation of the cDNA display library and *in vitro* selection against the SARS-CoV2 N protein was performed as previously reported (Haga et al., 2021), and N10 was subsequently obtained via NGS analysis. *In vitro* selection against SARS-CoV-2 S1 protein was performed by changing the immobilization method of the target protein from immune plate to magnetic beads, and E9 was obtained via NGS analysis.

### Gene construction (Nb-FP fusion)

The K-874A, E9, or N10 gene was amplified using primers containing the 5’-*Bam*HI and 3’-*Eco*RI sites, and the restricted products were cloned into the *Bam*HI/*Eco*RI sites of pBS Coupler 4 (Shimozono and Miyawaki, 2008) to generate pBS/K-874A=, pBS/E9=, or pBS/N10=, respectively. ‘=’ denotes the “coupler linker,” a triple repeat of the amino acid linker Gly–Gly–Gly–Gly–Ser [(GGGGS)_3_].

The KikG gene was amplified using primers containing the 5’-*Hin*dIII and 3’-*Sal*I sites, and the restricted product was cloned in-frame into the *Hin*dIII/*Sal*I sites of pBS/K-874A=, pBS/E9=, and pBS/N10= to generate pBS/K-874A=KikG, pBS/E9=KikG, and pBS/N10=KikG, respectively. The Azalea gene was amplified using primers containing the 5’-*Hin*dIII and 3’-*Sal*I sites, and the restricted product was cloned in-frame into the *Hin*dIII/*Sal*I sites of pBS/K-874A=, pBS/E9=, and pBS/N10= to generate pBS/K-874A=Azalea, pBS/E9=Azalea, and pBS/N10=Azalea, respectively. The EGFP or Achilles gene was amplified using primers containing the 5’-*Hin*dIII and 3’-*Sal*I sites, and the restricted product was cloned in-frame into the *Hin*dIII/*Sal*I sites of pBS/K-874A= to generate pBS/K-874A=EGFP or pBS/K-874A=Achilles, respectively.

In parallel, pRSET_B_ was engineered to have a *Sal*I site instead of a *Hin*dIII site.

The resultant plasmid was named pRSET_B_(*S*). The DNA fragments encoding K-874A=KikG, E9=KikG, N10=KikG, K-874A=Azalea, E9=Azalea, N10=Azalea, K-874A=EGFP, and K-874A=Achilles were cloned into the *Bam*HI/*Sal*I sites of pRSET_B_(*S*) to generate pRSET_B_(*S*)/K-874A=KikG, pRSET_B_(*S*)/E9=KikG, pRSET_B_(*S*)/N10=KikG, pRSET_B_(*S*)/K-874A=Azalea, pRSET_B_(*S*)/E9=Azalea, pRSET_B_(*S*)/N10=Azalea, pRSET_B_(*S*)/K-874A=EGFP, and pRSET_B_(*S*)/K-874A=Achilles, respectively.

### Visualizing SARS-CoV-2 spike protein in infected cultured cells

Infection experiments were conducted within a biosafety cabinet class II type B2 inside a biosafety level 3 laboratory. SARS-CoV-2 KUH003 strain (DDBJ accession number LC630936) was isolated from a COVID-19 patient hospitalized at Kitasato University Hospital (Haga et al., 2021). SARS-CoV-2 QK002 (Pango type; B.1.1.7, hCoV-19/Japan/QK002/2020, GISAID accession ID:EPI_ISL_768526) was used as the Alpha variant. SARS-CoV-2 TY8-612 (Pango type; B.1.351, hCoV-19/Japan/TY8-612/2021, GISAID accession ID:EPI_ISL_1123289) was used as the Beta variant. SARS-CoV-2 TY7-501 (Pango type; P.1, hCoV-19/Japan/TY7-501/2021, GISAID accession ID: EPI_ISL_833366) was used as the Gamma variant. These three variants were kindly provided by the National Institute of Infectious Diseases (NIID). VeroE6/TMPRSS2 cells were purchased from the Japanese Collection of Research Bioresources (JCRB) Cell Bank. Cells were fixed with cold methanol for 20 min and then incubated in Blocking solution I [phosphate-buffered saline (PBS) containing 3% BSA and 1% Triton X-100] for 60 min at room temperature (RT). After washing in PBS three times, the cells were reacted with 10 μg/mL fluorescent Nbs in Blocking solution I at RT for 60 min. In addition, nuclear staining was performed using DAPI (Dojin-Kagaku, 340-07971) (final conc. 1.0 μg/mL) at RT in PBS for 5 min.

Wide-field images were acquired using an inverted microscope (IX-83, Olympus) equipped with a lamp (X-cite XYLIS, EXCELITAS), a 20× dry objective (UPLXAPO, NA 0.8), and a CMOS camera (ORCA-Fusion, Hamamatsu Photonics). A U-FBNA filter cube (Olympus) was used to observe KikG, EGFP, and Achilles fluorescence. A U-FGNA filter cube (Olympus) was used to observe AzaleaB5 fluorescence. A U-FUNA filter cube (Olympus) was used to observe DAPI fluorescence. Image acquisition and analysis were carried out with cellSens Dimension (Olympus).

### Specimen collection

This study was approved by the ethics committee of Keio University (approval number: 20210081) and was conducted in accordance with the Declaration of Helsinki and Title 45, U.S. Code of Federal Regulations, Part 46, Protection of Human Subjects, effective December 13, 2001. All patients provided written informed consent.

### PCR measurement

SARS-CoV-2 RNA in saliva was detected using a rapid RNA extraction-free RT-qPCR kit (SARS-CoV-2 Direct Detection RT-qPCR Kit; Takara Bio, Kusatsu, Japan). This kit (approved by the Pharmaceuticals and Medical Devices Agency of Japan on October 27, 2020) used the primer sets for the amplification of two regions of the nucleocapsid gene, namely N1 and N2, as well as a human internal control gene. After inactivation by Solution A, samples were subjected to PCR measurement using the QuantStudio 5 Real-Time PCR System (Thermo Fisher Scientific, Waltham, MA, USA). After cDNA synthesis at 52°C for 5 min and denaturation at 95°C for 10 s, thermal cycling was performed (45 cycles of denaturation at 95°C for 5 s and annealing/extension at 60°C for 30 s). A sample was considered “positive” when amplification of the target region (N1 or N2 gene) was detected with a Ct value below 40.

### Oral cavity–derived samples for fluorescence microscopy observation

Each sample (SS or AS(OS)) (50 μL) in a 1.5 mL Eppendorf tube was combined with a 19-fold volume (950 μL) of 4% (wt/vol) PFA/PBS for fixation at RT overnight. After centrifugation at 35,000 ×g for 10 min, the precipitate was suspended with 50 μL PBS and stored at 4◻ until observation. An aliquot (7.5–10 μL) of the suspension was transferred into a 1.5-mL Eppendorf tube and mixed with 1 mL PBS. After centrifugation at 35,000 ×g at 8◻ for 10 min, 10 μL of a 99.5% ethanol solution (Fuji Film-Wako, 057-00456) was added to the precipitate. Dispersion of cells in approximately 70% ethanol solution was achieved by gentle tapping. The cell suspension was transferred with a plastic pipette onto the surface of a silicone-coated slide glass (Muto-Kagaku, 5116). Due to its low surface tension, the suspension spread quickly by itself. Most of the ethanol evaporated in a few minutes, which resulted in the generation of a sample of cells that were scattered homogenously in a circle with a diameter of several millimeters. The sample was enclosed with a water-repellent bank (Pap pen; Daido Sangyo). After washing with PBS twice, cells were treated with 0.7 mL Blocking solution II (salt solution containing 3% (wt/vol) BSA, 1% (wt/vol) Triton X-100, 50 mM PIPES, and 120 mM NaCl, pH 6.80) at RT for 30 min. The cells were incubated in 0.7 mL Blocking solution II containing a fluorescent Nb (10 μg/mL) at RT for 45 min. After washing with PBS twice, cells were fixed with 4% (wt/vol) PFA (pH 7.6) at RT for 10 min. After washing with PBS twice, cells were counterstained for nuclei in 0.7 mL PBS containing DAPI (Dojin-Kagaku, 340-07971) (final conc. 1.0 μg/mL) at RT for 5 min. After washing with PBS twice, the slide was mounted with a coverslip for observation by wide-field or confocal microscopy.

### Indirect immunocytochemistry for visualizing SARS-CoV-2 spike protein

Cells on a silane-coated slide glass (Muto-Kagaku, 5116) were treated with 0.7 mL PBS containing 0.2% (wt/vol) BSA, 0.1% (wt/vol) Triton X-100, and 17 μg/mL anti–SARS-CoV-2 spike mouse mAb (GeneTex, 1A9, GTX632604) at 37◻ for 30 min. After washing with PBS twice, cells were incubated in a 0.7 mL PBS containing 0.2% (wt/vol) BSA, 0.1% (wt/vol) Triton X-100, and 2.0 μg/mL Alexa Fluor 488–labeled goat anti–mouse IgG (H+L) (Thermo Fisher, A11001) or Alexa Fluor 546–labeled goat anti–mouse IgG (H+L)-F(ab’)2 fragment (Thermo Fisher, A11018) at 37◻ for 30 min.

### Visualizing squamous epithelial cells using an antibody against pan-cytokeratin

After reaction with a fluorescent Nb, cells were incubated in 0.7 mL PBS containing 0.1% (wt/vol) Triton X-100 and 1.25 μg/mL anti–pan-cytokeratin mouse mAb (AE1/AE3) (BioLegend, 914204) at RT for 60 min. After washing with PBS twice, cells were incubated in 0.7 mL PBS containing 0.1% (wt/vol) Triton X-100 and 2.0 μg/mL Alexa Fluor 633–labeled goat anti–mouse IgG (H+L)-F(ab’)2 fragment (Thermo Fisher/Invitrogen, A21053) at RT for 60 min.

### Delineating individual cells by immunostaining the plasma membrane

After reaction with a fluorescent Nb, cells were incubated in 0.7 mL PBS containing 0.1% (wt/vol) Triton X-100 and 3.3 μg/mL anti–pan-cadherin rabbit polyclonal antibody (abcam, ab16505) at RT for 60 min. After washing with PBS twice, cells were incubated in a 0.7 mL PBS containing 0.1% (wt/vol) Triton X-100 and 2.0 μg/mL Alexa Fluor 546–labeled donkey anti–rabbit IgG (H+L) antibody (Thermo Fisher/Invitrogen, A10040) at RT for 60 min.

### Confocal imaging

Saliva samples were imaged using an inverted laser scanning confocal microscopy system (Olympus FV3000) equipped with a 40× objective lens (UPLFLN40X, oil NA 1.30) and *z* drive. The *z*-step size was 1.5 μm. DAPI was excited by a 405-nm diode laser and fluorescence was detected within the wavelength range of 430–470 nm. KikG was excited by a 488-nm diode laser and fluorescence was detected within the wavelength range of 500–540 nm. Alexa Fluor 546 and Azalea were excited by a 561-nm diode laser and fluorescence was detected within the wavelength range of 570–611 nm. Alexa Fluor 633 was excited by a 640-nm diode laser and fluorescence was detected within the wavelength range of 650–750 nm. The size of the confocal aperture was ∼2 Airy disks. Multiple (3–6 × 3–6) *xy* images were tiled using the stitch function of FV3000 at each *z* position.

### Lyophilization

Immediately after purification, 0.1-mL aliquots of E9=KikG were made and frozen in liquid nitrogen. An aliquot was subjected to lyophilization using a freezing dryer (FDU-83, EYELA). The sample was reconstituted with 0.1 mL water.

### Quantification of SAES-CoV-2 infection

First, multiple fields of view (FOVs) containing observed cells were randomly selected. Second, individual cells were manually delineated using the “Freehand selections” tool (ImageJ) in each FOV. Third, the average intensity and the texture of the E9=KikG fluorescence were extracted using a customized program written using C++ and OpenCV 3.4.1 (https://opencv.org). Finally, labeling of the analyzed cells was classified as strong, sparse, or none using k-means with Origin Pro 2020b.

