## Supplementary Figures for "Antigen testing for COVID-19 using image-based assessment of oral specimens"

**Supplementary information**

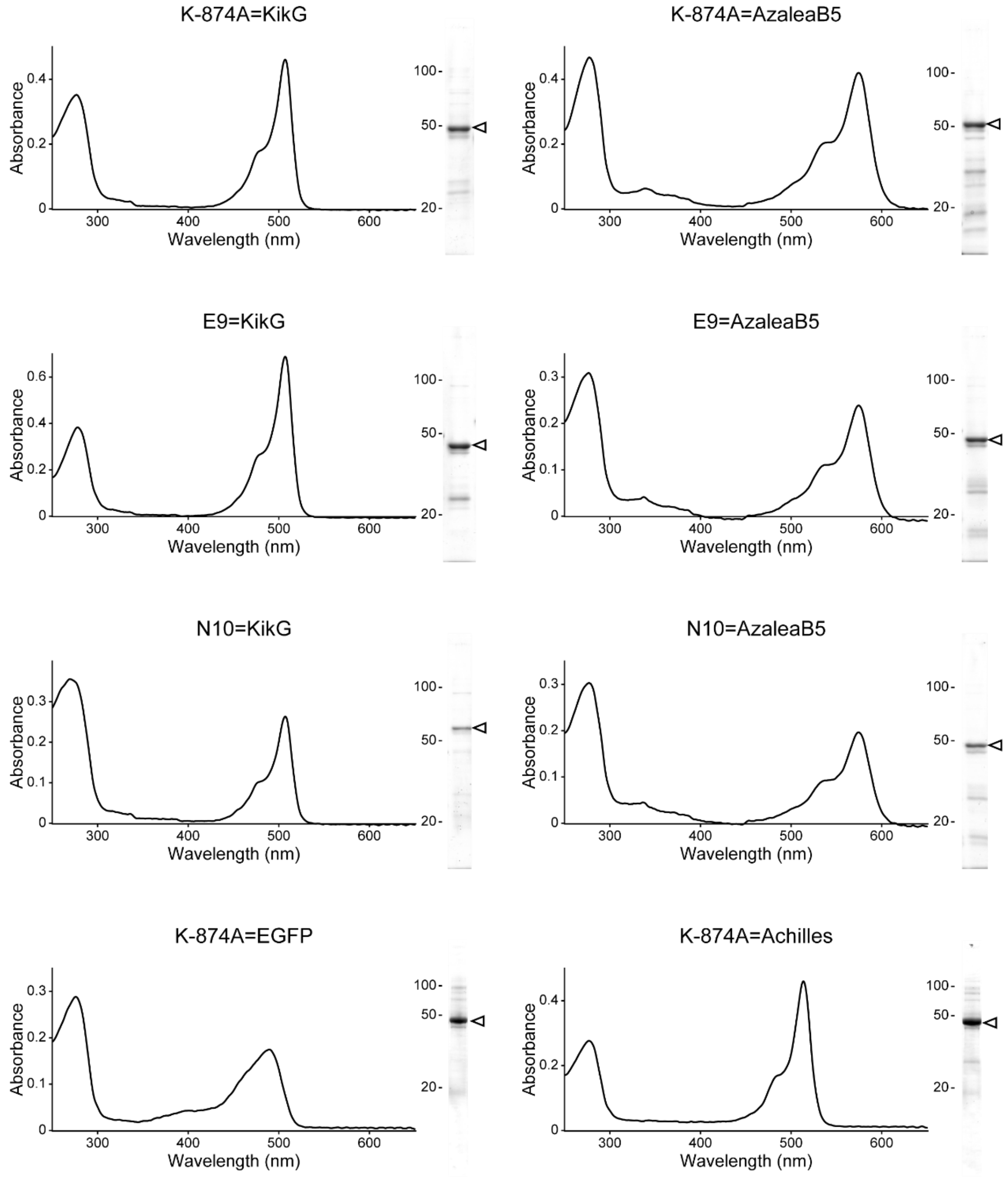


**Fig. S1**

Molecular and spectroscopic diagnosis of fluorescent Nbs (Nb-FP fusion proteins) used in this study. *left,* Absorption spectrum. *right,* Coomassie Brilliant Blue staining for the visualization of the purified protein separated by SDS-PAGE. The migration position is indicated by an arrowhead.

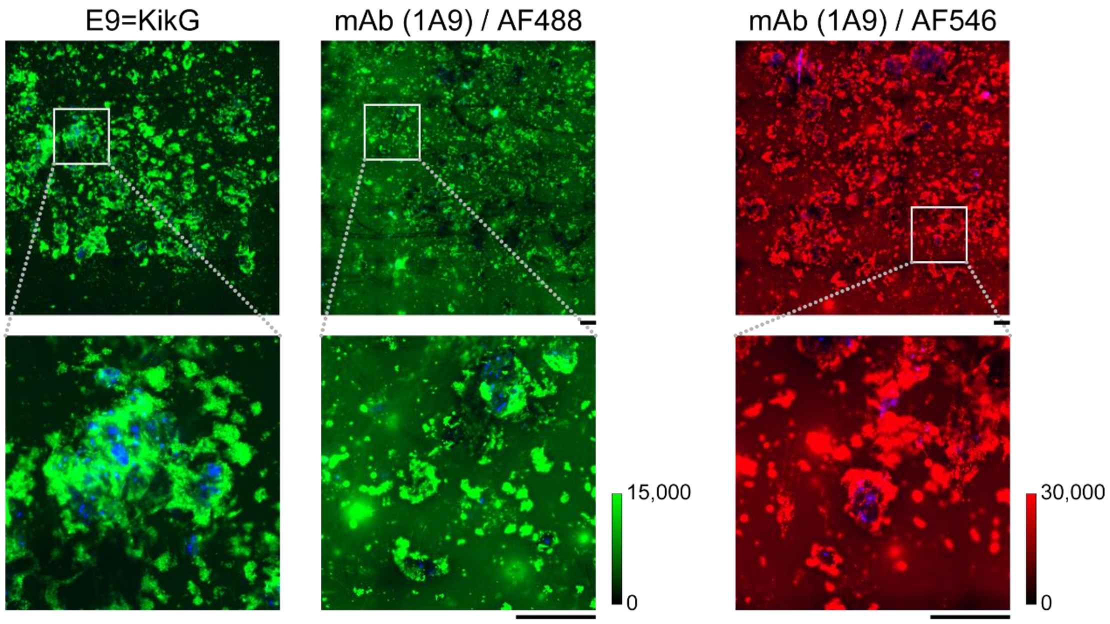


**Fig. S2**

Comparison of SNRs observed using E9=KikG versus a conventional immunocytochemical method with the 1A9 mAb. In the latter approach, secondary antibodies conjugated with Alexa Fluor 488 and Alexa Fluor 546 were used. Nuclei were counterstained with DAPI (blue). Fluorescence images were acquired by wide-field microscopy. Representative images of the #DP6 specimen are shown. The green and red scales indicate the lowest and highest image intensities. Scale bars, 0.2 mm.

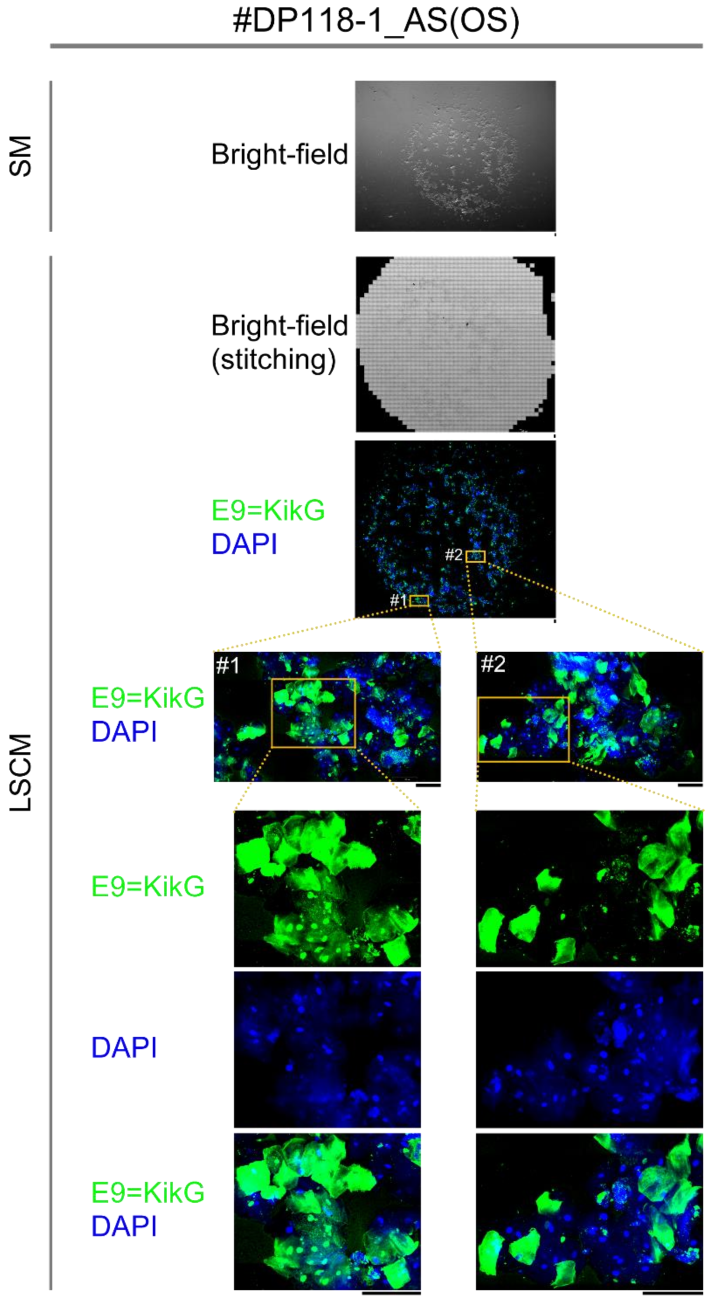


**Fig. S3**

Comprehensive dual-color image data for cells obtained from 7.5 μL of an AS(OS) specimen (#DP118-1_AS(OS)) taken from a COVID-19 patient (DP118) in the early stage of infection. To visualize nuclei (DAPI) and the SARS-CoV-2 spike protein (E9=KikG), the magnification was gradually increased while examining each region of interest. Scale bars, 0.1 mm. SM: stereomicroscope. LSCM: laser-scanning confocal microscope.

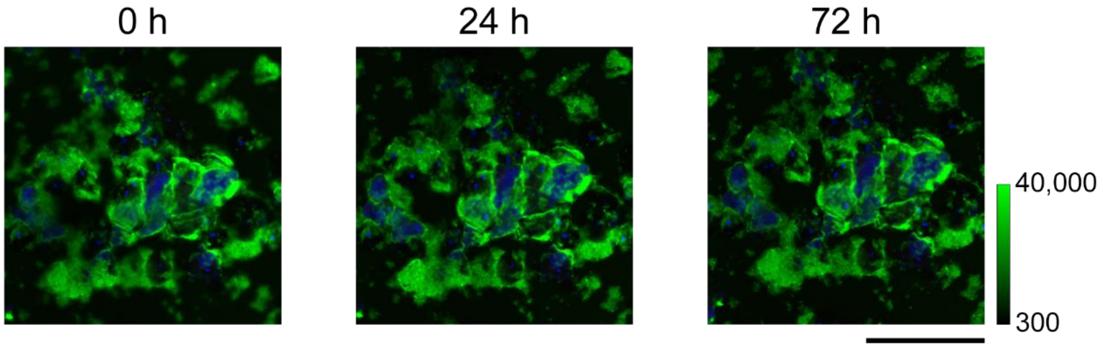


**Fig. S4**

Stability of E9=KikG immunosignals on an SS specimen (#DP50, Ct = 21.696) from a COVID-19 patient (DP50). Immediately after encapsulation with a coverslip, an area immunostained for E9=KikG (green) and DAPI (blue) was imaged by wide-field microscopy (0 h). The area was imaged again 24 h and 72 h later under the same optical conditions. The green scale indicates the lowest and highest image intensities. Scale bar, 0.2 mm.

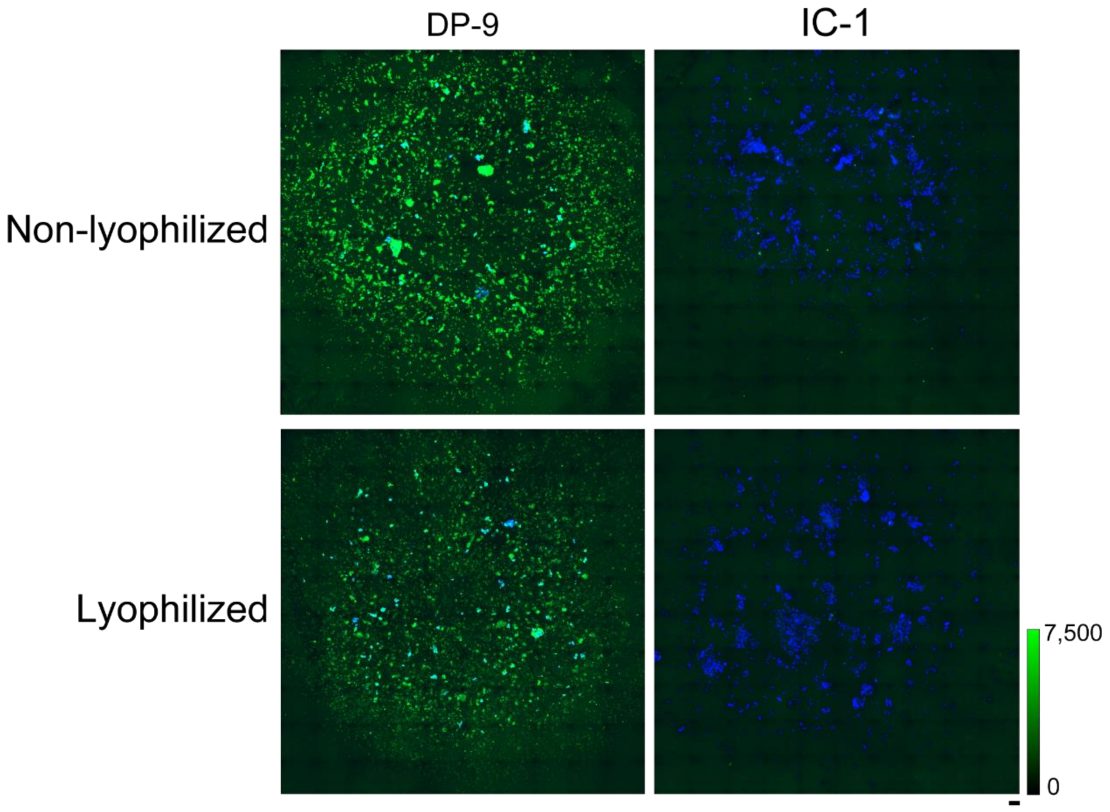


**Fig. S5**

Resistance of E9=KikG to lyophilization. After purification, dozens of aliquots of E9=KikG were prepared and frozen for storage. An aliquot was used to lyophilize the reagent. Scale bar, 0.2 mm.

*left,* Two smears derived from the AS(OS) specimen of a COVID-19 patient (DP9, Ct = 31.241) (see Fig. 3b) were immunoreacted with non-lyophilized (top) and lyophilized (bottom) reagents at the same concentration (10 μg/mL).

*right,* Two smears derived from the AS(OS) specimen of a healthy person (IC1, Ct > 40) (see Fig. 3b) were immunoreacted with non-lyophilized (top) and lyophilized (bottom) reagents at the same concentration (10 μg/mL).
